# Identifying Core Global Mental Health Professional Competencies: A Multi-Sectoral Perspective

**DOI:** 10.1101/2023.11.10.23298372

**Authors:** Dimitar Karadzhov, Joanne Lee, George Hatton, Ross White, Laura Sharp, Abdul Jalloh, Julie Langan Martin

## Abstract

Concerned with sustainably alleviating mental distress and promoting the right to health worldwide, global mental health (GMH) is practised across various contexts spanning the humanitarian-development-peace nexus. The inherently intersectoral and multidisciplinary nature of GMH calls for competency frameworks and training programmes that embody diversity, decolonisation and multiprofessionalism. Existing competency frameworks have failed to capture the multi-sectoral, inter-professional nature of contemporary GMH practice. In response to these needs, a qualitative content analysis of relevant job advertisements was conducted to distil a comprehensive set of professional competencies in contemporary GMH practice. Approximately 200 distinct skills and competencies were extracted from 70 job advertisements and organised into four meta-dimensions: ‘*skills*’, ‘*sector*’, ‘*self*’ and ‘*subject*’. The first known systematic attempt at a multi-sectoral GMH competency framework, it offers a springboard for exploring vital yet overlooked professional competencies such as resilience, self-reflection, political skills and entrepreneurialism. On this basis, recommendations for building a competent, agile and social justice-oriented GMH workforce with diversified and future-proof skillsets are proposed. The framework can also inform inter-professional training and curriculum design, and capacity-building initiatives aimed at early-career professional development, particularly in low- and middle-income settings.

**Impact Statement:** Professional competency frameworks play an important role in the education, training, employability and continuous professional development of the diverse – multi-disciplinary and multi-professional – global mental health workforce. To reflect this diversity, a novel, multi-sectoral global mental health competency framework was developed from a job market analysis and a stakeholder consultation. This framework encompasses a range of job families such as advocacy, policy, service delivery, programme management, capacity development and research and teaching. As such, it is applicable across geographical settings, career stages and global mental health-related job titles. Far from being a definitive list, the framework highlights the immense variety of interpersonal, technical, cognitive and knowledge-based competencies demanded from employers across sectors and roles. Alongside the well-recognised, quintessential competencies such as collaboration, cultural sensitivity, integrity and intervention delivery, educators, trainers, managers and other leaders should develop trainees’ and professionals’ resilience and adaptability; creativity and curiosity; and entrepreneurial and reflective skills. The professional development tool documented in this article can foster inter-professional mobility and education, together with the design of courses and curricula that are aligned with employer needs and contemporary challenges. Ultimately, the framework is designed to trigger reflection and professional skills analysis, and inspire lifelong learning.

## Introduction

Global mental health (GMH) is a professionally diverse field of practice spanning multiple sectors and roles (Collins, 2020). It is practised across humanitarian, development and peace settings (World Health Organization, 2013, 2021a). It is a nascent field, which is continuously redefining itself amidst ongoing debates about its scope, priorities and fundamental values (Rajabzadeh et al., 2021; Collins, 2020). Because GMH is characterised by a multivocality of disciplines, epistemologies, cultural perspectives and stakeholders, it defies easy categorisation (White et al., 2017). It eschews an essentialist view of mental disorders and well-being, and instead centres on context, collaboration, empowerment, humility and power-shifting in its pursuit to sustainably alleviate mental distress and promote the right to health worldwide. This expansiveness inherent to GMH practice generates distinct challenges to creating comprehensive training for future and current professionals in this field (Buzza et al., 2018; Acharya et al., 2024). As Collins (2020, p. 265) argued, ‘[g*]lobal mental health recognizes a vastly interconnected world and values nurturing that interconnectedness for solving difficult problems through a diversity of perspectives*.’. We hereby propose that systematic, concerted efforts to bolster professional development in this field must embody this interconnectedness - among cultures, positionalities, disciplines, sectors and skillsets.

### The Case for a Multi-sectoral, Multi-professional GMH Competency Framework

The deeply intersectoral nature of GMH calls for competency frameworks and training programmes that embody diversity and multiprofessionalism (Di Ruggiero, 2022). Rather than narrowly viewed as a healthcare issue, GMH has also been construed as a human rights and a development issue (White et al., 2016). GMH is practised across contexts with a significant presence within humanitarian and emergency settings. Roles often embrace the composite approach of *‘mental health and psychosocial support*’ (MHPSS) which is underpinned by inter- and multidisciplinary theoretical and conceptual frameworks and guidelines (e.g. Interagency Standing Committee, 2007). Correspondingly, there is a need for a resilient, collaborative and adaptive GMH workforce that can fulfil the promise of intersectoral and multi-stakeholder collaboration in a range of rapidly evolving contexts and address its Grand Challenges (Collins, 2020; Trowbridge et al., 2022).

One complicating factor in considering the variety of competencies across disciplines relates to the differences in how well established specific professional roles are in a global context. For instance, as identified within the World Health Organization Atlas 2020 (WHO, 2021b), mental health professionals may not have the same profile or presence in mental health across countries and regions; for example, specialised mental health nurses, occupational therapists, and speech therapists (WHO, 2022; World Federation of Occupational Therapists, 2022). A multitude of other relevant professional roles and the associated competencies may be comparatively under-represented - for instance, in the areas of capacity development, programme evaluation, policy, technical assistance, and advocacy. As Acharya and colleagues (2024, p. 88) note, ‘*GMH competencies will often go beyond clinical domains and incorporate health systems, leadership, advocacy, social sciences, education and training, cross-cultural navigation* […]’. This underscores the need for a comprehensive GMH competency framework that reflects this diversity and complexity of potential roles and responsibilities (Ng et al., 2016).

The international development funding landscape testifies to the interrelatedness between various mental health and non-mental health activities and roles (Liese et al., 2019). As shown in Liese and colleagues’ (2019) review of development assistance for health from all Organisation for Economic Co-operation and Development (OECD) sectors, funding was considerably less common for ‘pure’ mental health projects compared to projects related to other sectors in which mental health was one of several components. This demonstrates the need for GMH professionals to be able to competently contribute to intersectoral initiatives. This, in turns, requires strong, diversified and transferable professional skillsets.

Accordingly, recommendations have been made to enhance GMH professional training via ‘[…] *inter-professional and trans-professional education that breaks down professional silos and enhances collaboration* […] (Fricchione et al., 2012, p. 53). The ultimate goal, as Fricchione and colleagues (2012, p. 53) argue, should be to instil the skills, confidence and flexibility for professionals to access and scrutinise *‘global knowledge and experience*’ and apply them in solving ‘*local challenges*’. Relatedly, Fernando (2017) highlights that decolonising the GMH curricula should be prioritised. This may be especially important for students from medical and nursing backgrounds, since it has been recognised how mental health and mental ill health are taught within curricula has been heavily shaped by Eurocentric paradigms (Bracken et al., 2021). As such, this decolonisation process should increase student awareness of issues of discrimination and inequality within psychiatry and psychiatric services, that vary across localities.

Several recent endeavours have attempted to develop GMH competencies by either modifying related competency frameworks from adjacent disciplines (such as psychology), or by creating bespoke frameworks or lists of competencies (Khoury & De Castro Pecanha, 2023). Notably, most of these have been focused on psychological service delivery (Buzza et al., 2018; International Association of Applied Psychology & International Union of Psychological Science, 2016; World Health Organization and UNICEF, 2022), and (early-career) researchers (Merritt et al., 2019; Thornicroft et al., 2012; Collins and Pringle, 2016; Ng et al., 2016), and on specific interventions and settings such as humanitarian contexts (e.g. IFRC, 2016).

Relatedly, several interprofessional competency frameworks for *global health* have been put forward in recent years (e.g. Jogerst et al., 2015; Rowthorn and Olsen, 2014; Sawleshwarkar and Negin, 2017). While capturing the breadth of professional activities, skills and knowledge domains, such frameworks have said relatively little about the less quantifiable personal qualities and attributes that boost professional prospects such as resilience, determination, perseverance and curiosity. Moreover, there are concerns that competency lists developed by experts from curriculum reviews and other methods may not reflect the complexity and dynamism of contemporary professional practice (von Treuer and Reynolds, 2017).

To our knowledge, however, no unifying multi-sectoral GMH competency framework exists to date. A multi-sectoral, multi-professional GMH competency framework can strengthen work-related capacity of people at different stages in their careers, including early-career professionals and university students, as well as foster professionals’ international and interprofessional mobility and transition from nationalistic models of practice. In addition, it can empower educators, trainers and supervisors globally to better prepare students, trainees and mentees for the demands of contemporary roles and work settings (Acharya et al., 2024). Finally, such a tool can advance the discourse on the scope and remit of GMH (Rajabzadeh et al., 2021). Indeed, the systematic identification of guiding values and competencies is important for firmly establishing nascent disciplines such as GMH, and supporting the development of relevant professional identities (Salm et al., 2021).

### Understanding Employer Needs

In contrast to traditional methodologies of competency framework development such as subject expert consultations and desk-based curricular reviews, the analysis of job advertisements arguably offers a more objective, up-to-date and comprehensive overview of in-demand competencies (Brown et al., 2018; Keralis et al., 2018). It has been used successfully in fields such as global health to gauge employer expectations and boost sector awareness (Brown et al., 2018; Keralis et al., 2018).

### Research Aim and Methods

This study sought to identify a comprehensive set of in-demand GMH-related professional skills and competencies as indicated in relevant job descriptions and person specifications, across sectors and professional roles. To this end, a scoping job market analysis was carried out between September 2022 and January 2023, as part of a learning and teaching development project in a UK postgraduate taught Global Mental Health Master’s Degree programme.

Qualitative content analysis (QCA) was applied to extract discreet competencies from the job advertisements. QCA is a systematic and transparent method for parsing textual data into distinct entities and generating concise informative summaries in the form of conceptual categories, systems or maps (Elo and Kyngäs, 2008, p. 108). We adopted an inclusive definition of competencies as encompassing ‘*an interplay of knowledge, capacities and skills, motives and affective dispositions*’ (Rieckmann, 2012, p. 129). Consistent with the World Health Organization (2017, p. ix), we recognise competencies are not fixed but ‘*dynamic and contextual*’. This is a qualitative descriptive study underpinned by a *factist perspective*, whereby data are treated as ‘*more or less accurate and truthful indexes of the reality out there*’ (Vaismoradi et al., 2013, p. 400, citing Sandelowski, 2010).

### Data Collection

We used purposive sampling techniques, which aim to maximise the sample’s richness, diversity and informativeness, while not claiming statistical generalisability (Suri, 2011). Job listings with rich descriptions of roles and candidate profiles were given preference, together with those reflecting common and feasible career paths for graduates (*intensity sampling*; Suri, 2011). Job listings for a wide range of GMH-related roles and sectors were selected (*maximum variation*; Suri, 2011). The following jobs sites were searched: LinkedIn (www.linkedin.com/); UNJobs (https://unjobs.org); Charity Job (https://www.charityjob.co.uk); Rethink Mental Illness (https://www.rethink.org/); NGO Jobs in Africa (https://ngojobsinafrica.com/); Mind (https://www.mind.org.uk/about-us/working-for-us/local-mind-jobs/); Mental Health Innovation Network (https://www.mhinnovation.net/forums/vacancies-fellowships); Ghana Current Jobs (https://www.ghanacurrentjobs.com/); NHS Jobs (https://www.jobs.nhs.uk/); https://www.jobs.ac.uk/; and https://uk.indeed.com/. Initially, 50 job advertisements were selected, and sorted into job families. Then, another 20 were selected to add to any underrepresented job families in the initial sample. The final sample size of 70 was determined by the saturation observed – analysing additional advertisements did not yield new codes or competencies (Morse, 2015).

To track and ensure a multi-sectoral scope, the researchers categorised the 70 advertisements into several job families: advocacy (7); capacity development (8); policy (7); programme implementation, management and evaluation (12); research (15); clinical, psychological and psychosocial service delivery (17); and teaching (4). Exemplary roles were Regional Scaling Coordinator; Advocacy Worker; Assistant Psychologist; Policy Officer; Clinical Psychologist; Research Officer; WHO Consultant; and others. The jobs were based in the United Kingdom (34); the African continent (17, including Ethiopia, Nigeria, Tanzania, Zanzibar, South Sudan, Senegal, Kenya, Rwanda, Uganda, Kuwait, Ghana and Mali); multiple countries (4); other European countries (3); the Americas (3); New Zealand (2); and remote or other (7). A wide range of employers and sectors were represented, including humanitarian aid organisations, charities and other non-profit organisations, government agencies, higher education institutions, international NGOs, and the private sector.

### Data Analysis

The advertisements were exported into the qualitative data analysis software programme, NVivo 12 (https://support.qsrinternational.com/s/), where the person specifications, main responsibilities, required qualifications and employer information were manually analysed using QCA by the first and third authors. The purposive sampling and QCA aimed to map the *range* of relevant competencies; therefore, frequencies were not calculated. First, the advertisements were read and re-read line-by-line, following which codes corresponding to individual competencies were ascribed to short phrases or sentences (Elo and Kyngäs, 2008). Then, the long list of initial codes was re-examined, and codes were grouped into sub-categories based on similarities. The sub-categories were then clustered into a smaller number of higher-level, more abstract meaning units called categories (See ‘Table 1’, for an example of the coding process). Collectively, the categories conveyed the meaning of all codes parsimoniously. The third author conducted the initial coding, after which the first author reviewed this initial analysis for logical consistency, carried out further analysis, as required, and instigated the latter, abstraction process. The two authors met frequently to discuss coding decisions and resolve any discrepancies (Graneheim and Lundman, 2004).

**Table 1.**
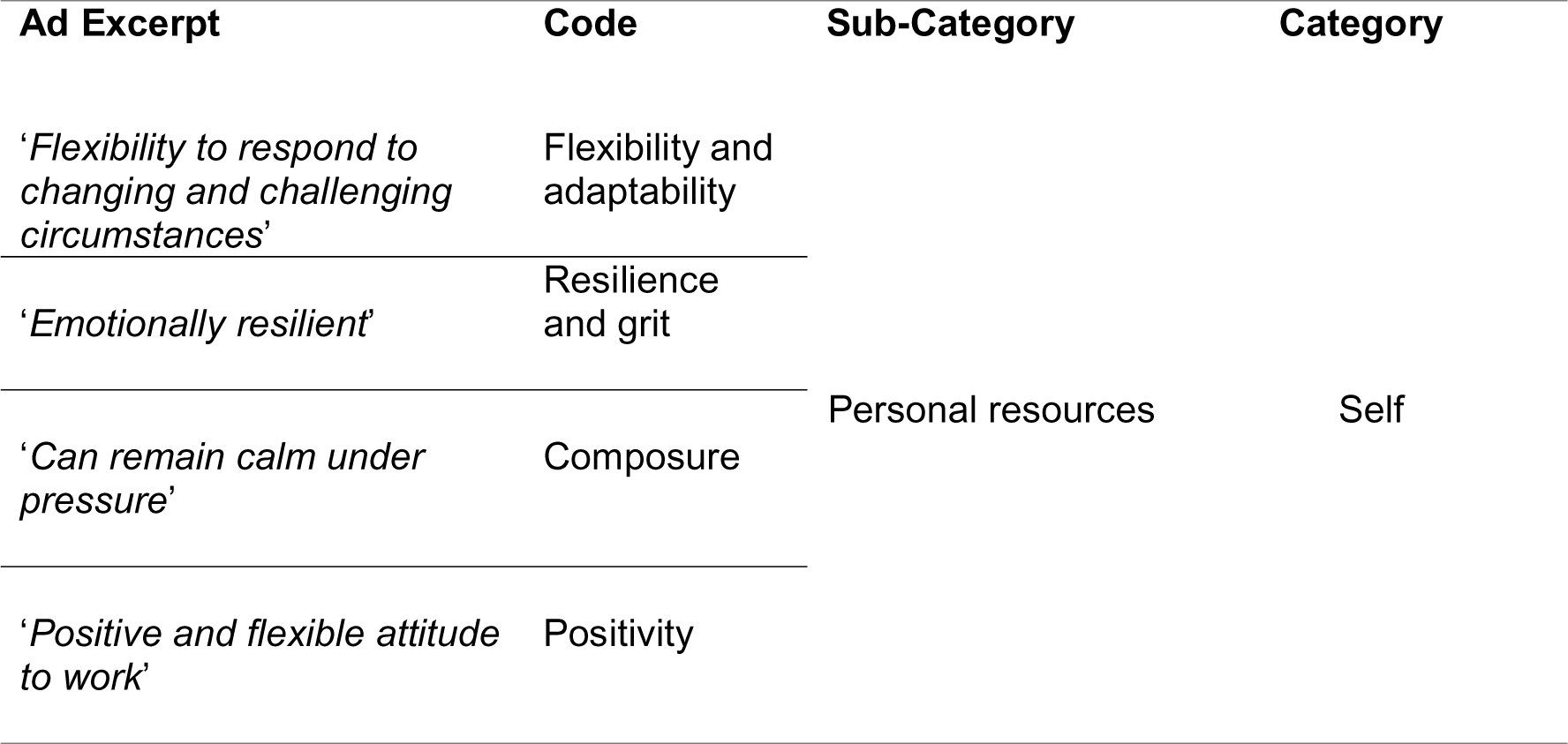
Example of the Qualitative Content Analysis Coding Process.

| Ad Excerpt | Code | Sub-Category | Category |
| --- | --- | --- | --- |
| *'Flexibility to respond to changing and challenging circumstances'* | Flexibility and adaptability | Personal resources | Self |
| *'Emotionally resilient'* | Resilience and grit |  |  |
| *'Can remain calm under pressure'* | Composure |  |  |
| *'Positive and flexible attitude to work'* | Positivity |  |  |

The framework was pilot-tested with 17 GMH professionals (seven experts including policy advisors, psychiatrists, clinical psychologists, lecturers, clinical managers and researchers from Ukraine, Sierra Leone, Uganda, Nigeria, Egypt and the United Kingdom, together with ten recent MSc Global Mental Health graduates). Their feedback helped identify and remove jargon and ambiguity from the framework, enhancing its international applicability.

## Results

Approximately 200 distinct skills and competencies were derived from the 70 job advertisements (See Supplementary Materials, for the full list). This reflected the fine-grained (line-by-line) QCA, together with the purposive, maximum variation sampling strategy. Four meta-level categories were found to reasonably accommodate the QCA categories and sub-categories (‘Figure 1’):

- *‘Self’* – enduring personal characteristics, abilities, and aptitudes;
- *‘Skills’* – transferable skills required across a wide range of professional settings, including technical and interpersonal skills);
- *‘Sector’* – skills, competencies, and experience required in specific roles and sectors;
- *‘Subject’* – working knowledge of theories, concepts, frameworks, and principles relevant to GMH research and practice.

**Figure 1.**
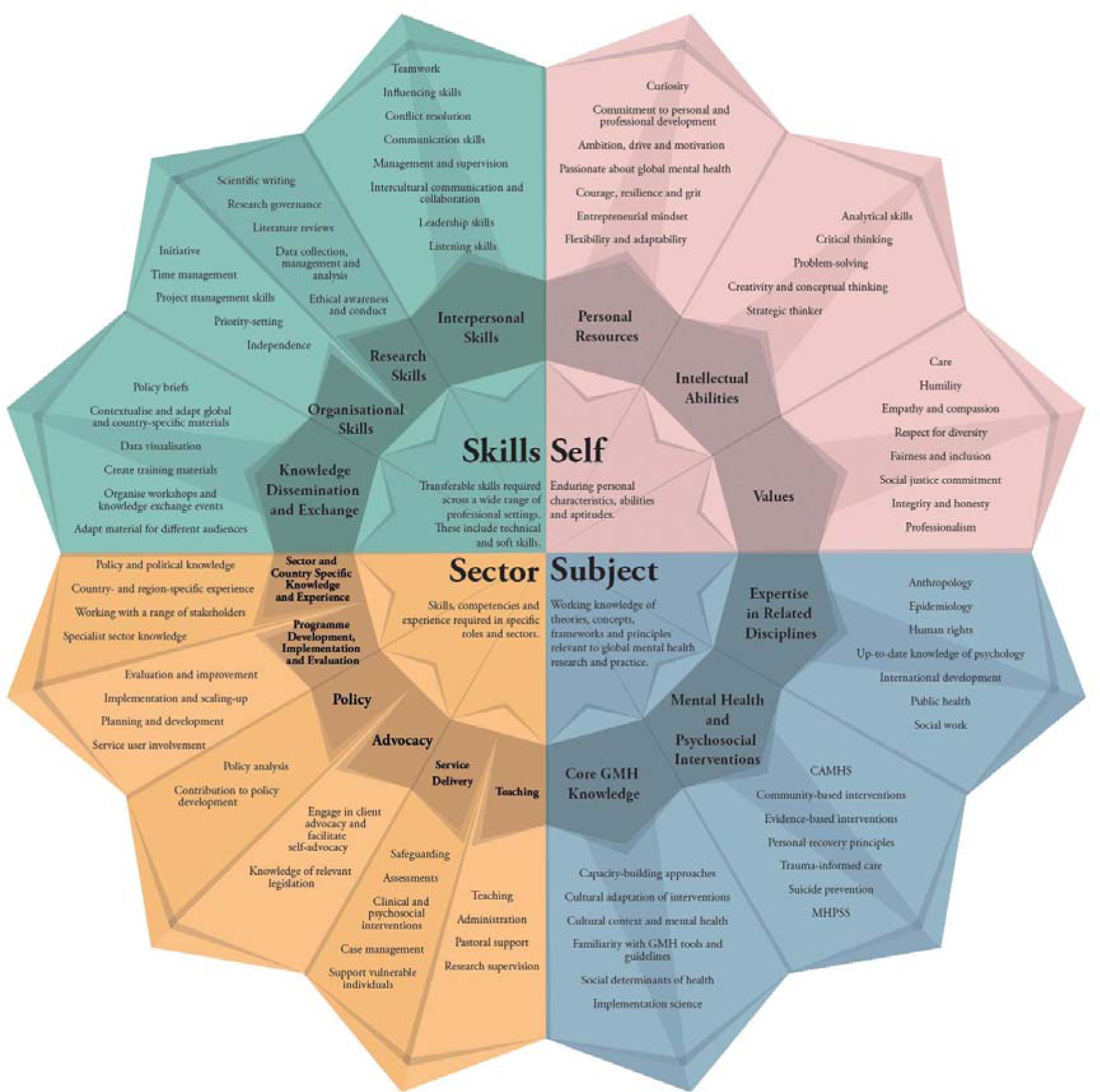
The ‘4S’ Multi-Sectoral Global Mental Health Competency Framework. *Note*. The list of competencies is not exhaustive. GMH - global mental health. CAMHS - child and adolescent mental health services. MHPSS - mental health and psychosocial support. **See Supplementary Files, for a higher-resolution file**.

### *Self*: Intellectual Abilities, Personal Resources, and Values

This category encompasses three sub-categories: intellectual abilities; personal resources; and values. *Intellectual abilities* are near-ubiquitous within job descriptions, and refer to cognitive abilities pertaining to analysing, synthesising and interpreting information and generating ideas and solutions. In addition to the quintessential skills of analytical and critical thinking, and problem-solving, employers placed a strong emphasis on creativity, idea generation and innovation. These were especially common in capacity development, service delivery and research roles. These attributes were often described as being open to change and ‘thriving’ in solving problems:

> ‘*Should thrive in solving problems and producing pragmatic solutions*’ (Regional Scaling Coordinator, War Child Holland)

> ‘*Research never goes exactly according to plan, and the Research Officer must approach the problems that will inevitably arise with patience and creativity* […] *Creativity and an ability to think outside the box to conceptualize projects and implementation strategies*’ (Research Officer - South Sudan, Forcier Consulting)

> ‘*Suggests creative improvements and better ways of working*’ (Research Coordinator, The MHPSS Collaborative)

> ‘*creative and proactive problem solver*’ (Social Worker - Ghana, International Justice Mission)The second sub-category, *personal resources*, is a highly heterogeneous cluster of traits and competencies that promote coping, resilience and thriving in the workplace (Kasler et al., 2017; See ‘Figure 1’). The most pervasive ones across sectors and roles were: (a) flexibility, adaptability and openness to change; (b) resilience and grit; and (c) ambition, drive and determination. Specifically, there was a strong focus on working effectively amidst changing, challenging and unclear circumstances. Notably, these were not reserved for humanitarian emergency jobs but were also required in service delivery roles in peace settings:

> ‘*Strong self-starter, able to take initiative and adapt to changing circumstances and priorities*’ (Psychological Counsellor - Ethiopia, Save the Children)

> ‘*Shifts tasks, roles and priorities to perform effectively under changing or unclear conditions*’ (Research Coordinator, The MHPSS Collaborative)

> ‘*Managing self: Displays grit, courage, resilience* […]’ (Senior Policy Advisor, Mental Health and Wellbeing Commission)Less commonly mentioned but noteworthy were self-discipline, curiosity, accountability, composure, courage, entrepreneurialism, among others – for example: ‘*entrepreneurial mindset*’ (Healthcare Partnerships & Service Integration - Kenya, Zipline); ‘*start-up mentality*’ (Lead Clinical Psychologist - Nigeria, Reliance Health); ‘*stand up and challenge decisions*’ (Advocacy Worker, Think Care Careers); ‘*we are courageous and speak up about what is important to people*’ (Senior Policy Advisor - New Zealand, Te Hiringa Mahara).

Finally, *values* are a sub-category that is distinct from intellectual abilities or traits and attributes promoting coping amidst change and adversity. This sub-category was derived from coding both the personal specifications and the employer information sections, and was prevalent across job families, including service delivery, advocacy and policy. A wide range of values were expected – including empathy, compassion and humility; respect, including respective for diversity; integrity and professionalism; social justice commitment; and fairness and inclusion:

> *‘Knowledge and commitment to anti-racist and inclusive practices*. [..] *Demonstrable commitment to upholding the rights of people who face disadvantage or discrimination*.’ (Independent Advocate, Gaddum Advocacy)

> ‘*Act in a way that acknowledges and recognizes peoples’ expressed beliefs, preferences and choices*.’ (Assistant Psychologist, NHS Wales)

### *Skills*: Interpersonal Skills, Research Skills, and Organisational Skills

This category accommodates the vast range of transferable skills – including *interpersonal skills, organisational skills* and *research skills* – that were the backbone of virtually all advertisements (See ‘Figure 1’). In addition to quintessential employability skills such as teamwork, time management, initiative and leadership, the analysis highlighted several more advanced and GMH-specific competencies, particularly *intercultural communication and collaboration*; *influencing skills*; *negotiation skills*; and *networking* and *relationship-building* within and across sectors:

> ‘*Excellent relationship building and influencing skills*’ (Senior Policy Officer - London, NHS)

> ‘*The role involves lobbying and developing understanding of the interventions and tools, organize and participate in adaption and contextualizing workshops and conducting online or face-to-face training with stakeholders and partners based in different countries across the region*’ (Regional Scaling Coordinator, War Child Holland)

> ‘*Work closely with the Psychological Intervention Specialist to conduct consultations with multi-sector stakeholders, community-based organizations, international NGOs*’ (Psychological Intervention Researcher, UNICEF)

> ‘*respectfully engage with partners and fellow staff members from different cultural backgrounds*’ (Research Officer - South Sudan, Forcier)Next, *research skills* represented a highly varied cluster comprising technical (e.g. data collection and analysis; information literacy); communication (e.g. scientific writing; grant proposals); and interpersonal and management skills. Importantly, research skills were found across job families, including service delivery, policy, capacity development and intervention evaluation.

Furthermore, several of the advertisements expected candidates to engage in various knowledge dissemination and exchange activities such as policy briefs, stakeholder dialogues, training materials and locally adapted guidelines:

> ‘*Preparation of accessible high quality reports on complex issues – for example policy positions, national consultations and member briefings*’ (Senior Policy Officer - London, NHS)

> ‘*Assist in the creation of training material and reports*’ (Intern - Peru, Innovations for Poverty Action)

> ‘*Adapt existing training packages to the current context and different needs*’ (MHPSS Technical Advisor, Red Cross)

### *Sector*: Sector- and Country-specific Competencies and Experience

The *sector* category houses competencies that are highly specific to the included job families - for instance, carrying out psychological assessments; cross-sectoral liaison; client advocacy; capacity and needs assessment; programme scale-up; and policy analysis and development (see ‘Figure 1’):

> ‘*Outstanding capacity to understand the country context, portfolio, and overall programmatic needs*’ (Mental Health Technical Advisor, International Rescue Committee)

> ‘*To work alongside and ensure active service user participation in all aspects of work, including design, implementation and monitoring of activities*’ (Mental Health Worker (Dual Diagnosis), Richmond Borough Mind)Pervasive across sectors and roles was the required ability to *work with a wide range of stakeholders*, including governmental and non-governmental agencies; professional groups; service-users; communities; and donors. This entailed demonstrable knowledge and understanding of key stakeholders at global and regional levels.

Relatedly, knowledge of relevant systems, legislation and policies was also commonly expected, including global frameworks and in-country policies:

> ‘*Knowledge of international policies, laws and mandates as pertaining to pesticides and suicides*’ (WHO Consultant)Finally, several jobs, mostly in capacity development, required experience of working in a LMIC, in war-affected regions, or a country different than one’s own.

### *Subject:* Core GMH Knowledge and Knowledge of Related Disciplines

Finally, this category represents the scientific- or knowledge-based competencies that were either essential or desirable candidate characteristics (See ‘Figure 1’). The analysis distinguishes between *core GMH knowledge* (for example, knowledge of scaling-up approaches and of cultural adaptation of interventions) and broader knowledge of *mental health and psychosocial interventions and theories*.

Partly due to the inclusive, multi-sectoral scope of the framework, employers commonly expected up-to-date knowledge of, or expertise in, related disciplines such as human rights, international development, social work, public health, anthropology and epidemiology.

## Discussion

This study aimed to scope the international GMH job market and extract and synthesise in-demand competencies from relevant job advertisements in order to develop a novel, multi-sectoral GMH competency framework. The framework accentuates the diversity of sectors, disciplines and transferable skills relevant to contemporary GMH practice, and highlights areas for workforce development (Ng et al., 2016; Fricchione et al., 2012). It is beyond the remit of this article to discuss and contextualise each competency identified in the current analysis; many of these have been discussed elsewhere (e.g. Merritt et al., 2019; Buzza et al., 2018; Thornicroft et al., 2012; Jogerst et al., 2015). Instead, the following sections lay out some of the most distinctive insights and practical implications of this scoping study.

### Values and Self-Reflection

The inclusion of values as a standalone competency dimension reflects not only their strong emphasis by employers but also GMH stakeholders’ assertions about the centrality of values to effective and equitable practice (Kohrt et al., 2016). The core values in the current framework (See ‘Figure 1’) cohere with other research on guiding principles and values in the field (Kohrt et al., 2016; Royal College of Psychiatrists, 2017). What has received significantly less attention is *how* to instil and nurture these values in GMH students and trainees. Crucially, GMH trainees and job candidates should be capable of articulating their core values, demonstrating their continuity in their personal and professional experiences, and evidencing a willingness to uphold these values in challenging circumstances. This requires, among other meta-competencies, strong self-reflection and self-analysis (Kohrt et al., 2016).

### Ubiquity of Research Skills

A noteworthy finding of the job market analysis was the presence of research skills in non-research positions such as policy, programme management, and psychological service delivery. Research skills training is essential to building capacity for GMH, particularly in LMICs (Wainberg et al., 2017; Okewole et al., 2020). Adoption of research competencies has been shown to boost personal and professional growth (Okewole et al., 2020), as well as increasing capacity for implementing evidence-based care (Thornicroft et al., 2012; Wainberg et al., 2017; Merritt et al., 2019). As Abu-Zaid (2014) shows, few medical students show interest in undergraduate research engagement as well as research-based careers mainly due to factors such as insufficient exposure to scientific research early in education, unwillingness to prolong medical training, personal preference, and failure to understand the importance of having research skills in general practice. Thus, building a positive attitude towards research remains a priority (Merritt et al., 2019). Merritt and colleagues (2019) highlight the value of improving research skills in those working in the GMH field in low- and middle-income countries to help rebalance historic underinvestment. The current job market analysis also underscores the need to scale up access to professional development programmes for GMH professionals focusing on knowledge-to-practice translation (Wainberg et al., 2017).

### Communication and Political Skills

While an array of communication skills have invariably featured in existing competency frameworks (Buzza et al., 2018; Merritt et al., 2019), the current job market analysis shines a light on a broader spectrum of communication and political skills, including some less commonly emphasised skills such as conflict resolution and influencing skills, together with policy and political knowledge and sensitivity. Multi-stakeholder collaboration, negotiation and consensus-building are indispensable in many GMH-related roles, particularly in the policy, advocacy, capacity-building and healthcare management sectors (Ng et al., 2016; Iemmi, 2022). Negotiation, specifically, has been highlighted as an emotionally demanding skill – hence its close links to leadership, emotional intelligence and resilience (Higazee and Gab Allah, 2022). This signals the need to embed such training in GMH programmes – for instance, in the form of simulation and role-play activities (Higazee and Gab Allah, 2022).

### Emphasis on Higher-Order Competencies

A distinctive finding of the current analysis – over and above existing frameworks – is the strong emphasis on dispositional and motivational characteristics such as creativity, strategic thinking, perseverance and curiosity. This tendency was also observed in Keralis et al.’s (2018) market analysis of global health jobs. Some of these competencies (for example, resilience, entrepreneurial mindset, courage, curiosity, perseverance, passion and adaptability) may be difficult to measure or evaluate, and have therefore remained under-investigated in the professional skills development literature (Jogerst et al., 2015; von Treuer and Reynolds, 2017). We nevertheless propose they remain important, and often integral, to effective professional activity in the GMH field.

Rowthorn and Olsen (2014) address this tension in their discussion of team skills competencies in global health education. Specifically, their roundtable expert participants underscored the importance of personal attributes for vocational success, including those that may be more difficult to teach or assess such as mindfulness, openness, vulnerability, sensitivity and self-discipline. The participants believed such non-cognitive attributes or dispositions should be integrated into disciplinary competency frameworks. As Jardim (2021, p. 1) argues, ‘[c]*ognitive and technical skills are not sufficient to face the professional challenges of the current digital and global world* […]’. Moreover, these personal attributes may serve as *meta-competencies* - ‘[…] *mindsets, comprising primarily of salient and relatively enduring attitude*lJ*values that have the potential to shape the growth of other competencies*.’ (Gonsalvez and Calvert, 2014, p. 205).

For educators and managers, this presents the challenge of designing appropriate approaches to fostering and assessing these competencies (Schleiff et al., 2020). Techniques such as scenario-based learning and peer feedback have shown promise in this area (Wroe et al., 2017). Many of the identified core attitudinal and technical competencies are likely best developed in the field. To allow for students’ and trainees’ exposure to authentic professional and cultural settings, multi-country partnerships, particularly between high-income and low- and middle-income countries, should be established (Marienfield et al., 2024).

### Resilience and Adaptability

The inclusion of resilience and adaptability in the current framework is not surprising given the challenging, resource-constraints contexts of much of GMH practice, together with the often-stigmatised, underfunded and neglected issues professionals in this field seek to redress. GMH professionals operate in ideologically and professionally contested and politically and economically unstable settings, and are therefore required to be adaptable, resilient and persevering; they are required to be path-finders (White et al., 2017; Dean et al., 2020). Most recently, the COVID-19 pandemic has demonstrated, and reinvigorated interest in, the importance of fostering resilience in vital professional groups such as healthcare and aid workers (Dean et al., 2020; Young et al., 2022). Fundamentally, the capacities for resilience and adaptability have been identified as core to sustainable development (Rieckmann, 2012; Brundiers et al., 2021).

As Matheson and colleagues (2016) note, it would be fruitful to explore the practices and characteristics of resilient professionals, and assess the extent to which those could be trained and taught. Among the resilience-promoting workplace interventions and behaviours suggested by Matheson et al.’s (2016) participants are: exposure to challenging situations; peer learning; and mindfulness. There is a strong case, therefore, that such practices should be embedded in GMH curricula. Developing a resilient workforce will require approaches that go beyond didactic, classroom learning. Learners and trainees should be provided with opportunities for experiential learning through internships, mentorship, site visits and engagement with communities of practice.

### Entrepreneurialism and GMH

Entrepreneurial skills and aptitudes (such as a start-up mentality, strategic thinking, perseverance, persuasion, optimism, generativity, drive and flexibility) also emerged as important. However, these have remained overlooked in existing competency frameworks. Acute global crises such as COVID-19, together with rapid technological advancements such as the rise of digital healthcare and artificial intelligence, have provided fertile ground for *global entrepreneurship*, including in LMICs (Mishra and Pandey, 2023). Against this backdrop, a relatively small but growing body of literature has testified to the significance of (social) entrepreneurship in realising the GMH agenda (Kidd et al., 2015).

We hereby argue that an entrepreneurial lens offers an opportunity to reinvigorate and enrich GMH training by increasing the focus on developing trainees’ creative, enterprising and leadership capabilities (Tang et al., 2018). We urge educators, trainers, managers and other leaders to explore ways to cultivate entrepreneurial skills in the GMH workforce. The importance of entrepreneurialism demonstrated here can inspire the wider use of collaborative and digitally-mediated learning modalities in GMH curricula – for example, by leveraging international partnerships, internships, mentorship and alumni involvement (Colombelli et al., 2022). Exposing students and trainees to social entrepreneurship role models is also beneficial. Entrepreneurial qualities are highly transferable, and will be relevant to those working in a range of GMH job families such as policy-making, capacity development and organisation management.

### Suggested Applications of the Framework

Altogether, the current framework can be used to guide GMH and allied professionals in identifying professional training priorities, and encourage lifelong learning (Okewole et al., 2020; Jogerst et al., 2015). It can also aid current students and trainees in career planning, including job applications, and in promoting self-awareness and reflection on professional and academic progress. For prospective students, it can be used to make GMH programmes more attractive by demonstrating the breadth and transferability of skills that can be acquired (Acharya et al., 2024). Moreover, it can aid educators in designing and institutionalising programmes aligned with the job market, including collaborative, interprofessional training programmes (Rowthorn & Olsen, 2014; Acharya et al., 2024; Okewole et al., 2020). Many of the competencies identified in the framework can directly inform creative authentic assessments and training opportunities, and equip trainees to undertake the decolonisation process.

### Limitations of the Framework and Future Directions

The framework discussed in this paper is limited in several ways. Fundamentally, although effort was made to include advertisements from different countries, the framework presents a Eurocentric interpretation of the professional skills required by the GMH workforce. It would be beneficial to further engage with the requirements in posts embedded in community practice across the globe. Currently there is a potential bias to employment opportunities created by the representatives of the Global North who may unintentionally prioritise Western values. Furthermore, the sample of job advertisements selected for its development (N = 70) is relatively small, and although a degree of saturation was reached, many regions and job roles have remained underrepresented; in particular, advertisements that do not use traditional mental health job titles but are within the GMH scope – for example, child protection and occupational therapy roles. Therefore, more extensive searches and more systematic validation approaches (using techniques such as Delphi) are recommended.

Moreover, the framework was initially designed to boost the employability of English-speaking GMH graduates in the UK, and due to its reliance on job advertisements as the data sources, one could argue it represents an *employability framework* more so than a competency framework. Further, larger-scale validation is recommended, particularly by engaging a wider range of professional groups and other stakeholders to maximise its inclusivity and relevance. Further work is also warranted into (a) translating the framework into training guides that can be adapted for different sectors, professional groups and cultural settings; and (b) instrumentalising the core competencies into learning objectives (Brundiers et al., 2021; Schleiff et al., 2020).

Finally, leadership, resilience, creativity, adaptability and other individual traits can also be viewed as emergent and relational properties of the organisational culture and structures and team dynamics (Masten and Motti-Stefanidi, 2020; Dean et al., 2020). We therefore encourage readers to reflect on the applicability of the Multi-Sectoral GMH Competency Framework at the organisational and team levels.

## Supporting information

Multi-Sectoral Global Mental Health Employability Framework

## Data Availability

All data produced in the present study are available upon reasonable request to the authors.

## Acknowledgements

We would like to sincerely thank the MSc Global Mental Health alumni and professionals who offered feedback on the competency framework, and Ian Hutchison, for the graphic design. We would also like to thank Ms. Mia Wilson, for the learning development support, and Ms. Fiona Stubbs (University of Glasgow), for the continued support with this project.

## References

Abu-Zaid A (2014) Research skills: the neglected competency in tomorrow’s 21st-century doctors. Perspectives on Medical Education 3(1), 63–65.

Acharya B, Buzza C, Guo J, Basnet M, Hung E and Van Dyke C (2024) Developing a global mental health training curriculum. In Global Mental Health Training and Practice, (ed. B. Acharya and AE. Becker), pp. 81–95. New York and Oxon: Routledge.

Bracken P, Fernando S, Alsaraf S, Creed M, Double D, Gilberthorpe T, Timimi S. (2021). Decolonising the medical curriculum: Psychiatry faces particular challenges. Anthropology & Medicine 28(4), 420–428. 10.1080/13648470.2021.1949892

Brown HA, Mulherin P, Ferrara WC, Humphrey ME, Vera A and Hall JW (2018) Using future employers’ expectations to inform global health fellowship curricula. Journal of Graduate Medical Education 10(5), 517–521. 10.4300/JGME-D-18-00348.1

Brundiers K, Barth M, Cebrián G, Cohen M, Diaz L, Doucette-Remington S, … and Zint M (2021) Key competencies in sustainability in higher education—toward an agreed-upon reference framework. Sustainability Science 16, 13–29. 10.1007/s11625-020-00838-2

Buzza C, Fiskin A, Campbell J, Guo J, Izenber, J, Kamholz B, … and Acharya B (2018) Competencies for global mental health: developing training objectives for a post-graduate fellowship for psychiatrists. Annals of Global Health 84(4), 717–726. doi: 10.29024/aogh.2382

Collins PY (2020) What is global mental health? World Psychiatry 19(3), 265. doi: 10.1002/wps.20728

Collins PY and Pringle BA (2016) Building a global mental health research workforce: perspectives from the National Institute of Mental Health. Acad Psychiatry 40, 723–726. 10.1007/s40596-015-0453-3

Colombelli A, Loccisano S, Panelli A, Pennisi OAM and Serraino F (2022) Entrepreneurship education: the effects of challenge-based learning on the entrepreneurial mindset of university students. Administrative Sciences 12(1), 10. 10.3390/admsci12010010

Dean L, Cooper J, Wurie H, Kollie K, Raven J, Tolhurst R … and Mansaray B (2020) Psychological resilience, fragility and the health workforce: lessons on pandemic preparedness from Liberia and Sierra Leone. BMJ Global Health 5(9), e002873. doi:10.1136/bmjgh-2020-002873

Di Ruggiero E (2022) Addressing mental health through intersectoral action in the context of COVID-19 and the 2030 Agenda for Sustainable Development. Global Health Promotion 29(3), 3–4. 10.1177/17579759221122710

Elo S and Kyngäs H (2008) The qualitative content analysis process. Journal of Advanced Nursing 62(1), 107–115. 10.1111/j.1365-2648.2007.04569.x

Fernando S (2017) Institutional Racism in Psychiatry and Clinical Psychology. London: Palgrave Macmillan.

Fricchione GL, Borba CP, Alem A, Shibr T, Carney JR and Henderson DC (2012) Capacity building in global mental health: professional training. Harvard Review of Psychiatry 20(1), 47–57. DOI: 10.3109/10673229.2012.655211

Gonsalvez CJ and Calvert FL (2014) CompetencylJbased models of supervision: principles and applications, promises and challenges. Australian Psychologist 49(4), 200–208. 10.1111/ap.12055

Graneheim UH and Lundman B (2004) Qualitative content analysis in nursing research: concepts, procedures and measures to achieve trustworthiness. Nurse Education Today 24, 105–112. doi: 10.1016/j.nedt.2003.10.001

Higazee MZA and Gab Allah AR (2022) The relationship between the political skills and negotiation behaviors of frontlJline nursing managers. Nursing Forum 57(6), 1240–1248. 10.1111/nuf.12772

IFRC (2016) Competency framework: psychosocial support delegates in emergencies. https://pscentre.org/wp-content/uploads/2018/10/Final-versionCompetency-Framework-September-16.pdf (accessed August 31 2023).

Iemmi V (2022) Establishing political priority for global mental health: a qualitative policy analysis. Health Policy and Planning 37(8), 1012–1024. 10.1093/heapol/czac046

International Association of Applied Psychology & International Union of Psychological Science (2016) International declaration of core competencies in professional psychology. https://www.iupsys.net/wp-content/uploads/2021/09/the-international-declaration-on-core-competences-in-professional-psychology-1.pdf (accessed August 31 2023).

Jardim J (2021) Entrepreneurial skills to be successful in the global and digital world: proposal for a frame of reference for entrepreneurial education. Education Sciences 11(7), 356. 10.3390/educsci11070356

Jogerst K, Callender B, Adams V, Evert J, Fields E, Hall T … and Wilson LL (2015) Identifying interprofessional global health competencies for 21st-century health professionals. Annals of Global Health 81(2), 239–247. 10.1016/j.aogh.2015.03.006

Kasler J, Zysberg L, and Harel, N (2017) Hopes for the future: demographic and personal resources associated with self-perceived employability and actual employment among senior year students. Journal of Education and Work 30(8), 881–892. 10.1080/13639080.2017.1352083

Keralis JM, Riggin-Pathak BL, Majeski T, Pathak BA, Foggia J, Cullinen KM … and West HS (2018) Mapping the global health employment market: an analysis of global health jobs. BMC Public Health 18, 1–9. 10.1186/s12889-018-5195-1

Khoury B and De Castro Pecanha V (2023) Transforming psychology education to include global mental health. Cambridge Prisms: Global Mental Health 10, E17. 10.1017/gmh.2023.11

Kidd SA, Kerman N, Cole D, Madan A, Muskat E, Raja S, Rallabandi S and McKenzie K (2015) Social entrepreneurship and mental health intervention: a literature review and scan of expert perspectives. Int J Ment Health Addiction 13, 776–787. 10.1007/s11469-015-9575-9

Kohrt BA, Marienfeld CB, Panter-Brick C, Tsai AC and Wainberg ML (2016) Global mental health: five areas for value-driven training innovation. Academic Psychiatry 40, 650–658. 10.1007/s40596-016-0504-4

Liese BH, Gribble RS and Wickremsinhe MN (2019) International funding for mental health: a review of the last decade. International Health 11(5), 361–369. 10.1093/inthealth/ihz040

Marienfeld C, Hu X, Yang Y, Liu, Z, Lasswell E and Rohrbaugh RM (2024) Educational partnerships: addressing challenges in meeting trainee goals with established or new global mental health educational programs. In Global Mental Health Training and Practice, (ed. B. Acharya and AE. Becker), pp. 135–144. New York and Oxon: Routledge. DOI: 10.4324/9781315160597-10

Masten AS and Motti-Stefanidi F (2020) Multisystem resilience for children and youth in disaster: reflections in the context of COVID-19. Adversity and Resilience Science 1(2), 95–106. 10.1007/s42844-020-00010-w

Matheson C, Robertson HD, Elliott AM, Iversen L and Murchie P (2016) Resilience of primary healthcare professionals working in challenging environments: a focus group study. British Journal of General Practice 66(648), e507–e515. DOI: 10.3399/bjgp16X685285

Merritt C, Jack H, Mangezi W, Chibanda D and Abas M (2019) Positioning for success: building capacity in academic competencies for early-career researchers in sub-Saharan Africa. Global Mental Health 6, E16. doi: 10.1017/gmh.2019.14

Mishra A and Pandey N (2023) Global entrepreneurship in healthcare: a systematic literature review and bibliometric analysis. Global Business and Organizational Excellence. DOI: 10.1002/joe.22193

Morse JM (2015) Data were saturated… Qualitative Health Research 25(5), 587–588. 10.1177/1049732315576699

Ng LC, Magidson JF, Hock RS, Joska JA, Fekadu A, Hanlon C … and Henderson DC (2016) Proposed training areas for global mental health researchers. Academic Psychiatry 40(4), 679–685. DOI: 10.1007/s40596-016-0518-y

Okewole H, Merritt C, Mangezi W, Mutiso V, Jack HE, Eley TC and Abas M (2020) Building career development skills for researchers: a qualitative study across four African countries. Annals of Global Health 86(1). PMID: 32322538

Rajabzadeh V, Burn E, Sajun SZ, Suzuki M, Bird VJ and Priebe S (2021) Understanding global mental health: a conceptual review. BMJ Global Health 6(3), e004631. 10.1136/bmjgh-2020-004631

Rieckmann M (2012) Future-oriented higher education: which key competencies should be fostered through university teaching and learning? Futures 44(2), 127–135. 10.1016/j.futures.2011.09.005

Rowthorn V and Olsen J (2014) All together now: developing a team skills competency domain for global health education. Journal of Law, Medicine & Ethics 42(4), 550–563. https://link.gale.com/apps/doc/A401904458/AONE?u=ustrath&sid=bookmark-AONE&xid=0be735cf

Royal College of Psychiatrists (2017) Core values for psychiatrists. https://www.rcpsych.ac.uk/docs/default-source/improving-care/better-mh-policy/college-reports/college-report-cr204.pdf?sfvrsn=5e4ff507_4 (accessed November 30, 2023).

Salm M, Ali M, Minihane M and Conrad P (2021) Defining global health: findings from a systematic review and thematic analysis of the literature. BMJ Global Health 6(6), e005292. doi:10.1136/bmjgh-2021-005292

Sandelowski M. (2010) What’s in a name? Qualitative description revisited. Research in Nursing & Health, 33(1), 77–84. 10.1002/nur.20362

Sawleshwarkar S and Negin J (2017) A review of global health competencies for postgraduate public health education. Frontiers in Public Health 5, 46. 10.3389/fpubh.2017.00046

Schleiff M, Hansoti B, Akridge A, Dolive C, Hausner D, Kalbarczyk A … and Bennett S (2020) Implementation of global health competencies: a scoping review on target audiences, levels, and pedagogy and assessment strategies. PLoS One 15(10), e0239917. 10.1371/journal.pone.0239917

Suri H (2011) Purposeful sampling in qualitative research synthesis. Qualitative Research Journal 11(2), 63–75. 10.3316/QRJ1102063

Tang C, Byrge C and Zhou J (2018) Creativity perspective on entrepreneurship. In The Palgrave Handbook of Multidisciplinary Perspectives on Entrepreneurship, (ed. R Turcan and N Fraser). Palgrave Macmillan, Cham. 10.1007/978-3-319-91611-8_5

Thornicroft G, Cooper S, Bortel TV, Kakuma R and Lund C (2012) Capacity building in global mental health research. Harvard Review of Psychiatry 20(1), 13–24. doi: 10.3109/10673229.2012.649117

Trowbridge J, Tan JY, Hussain S, Osman AEB and Di Ruggiero E (2022) Examining intersectoral action as an approach to implementing multistakeholder collaborations to achieve the sustainable development goals. International Journal of Public Health 67, 1604351. 10.3389/ijph.2022.1604351

von Treuer KM and Reynolds N (2017) A competency model of psychology practice: articulating complex skills and practices. Frontiers in Education 2, 54. doi: 10.3389/feduc.2017.00054

Vaismoradi M, Turunen, H and Bondas T (2013) Content analysis and thematic analysis: implications for conducting a qualitative descriptive study. Nursing & Health sciences, 15(3), 398–405. doi: 10.1111/nhs.12048

Wainberg ML, Scorza P, Shultz JM, Helpman L, Mootz JJ, Johnson KA … and Arbuckle MR (2017) Challenges and opportunities in global mental health: a research-to-practice perspective. Current Psychiatry Reports 19, 1–10. 10.1007/s11920-017-0780-z

White RG, Imperiale, MG and Perera E (2016) The Capabilities approach: fostering contexts for enhancing mental health and wellbeing across the globe. Globalization and Health 12(1), 1–10. 10.1186/s12992-016-0150-3

White RG, Orr DM, Read UM and Jain S (2017) Situating global mental health: sociocultural perspectives. In The Palgrave Handbook of Sociocultural Perspectives on Global Mental Health, (ed. RG White, S Jain, DMR Orr and UM Read), pp. 1–27. London: Springer Nature.

World Federation of Occupational Therapists (2022) Human Resources Project 2022: Global demographics of the occupational therapy profession. https://wfot.org/resources/occupational-therapy-human-resources-project-2022-numerical (accessed 22 October 2023)

World Health Organization (2013) Building back better: sustainable mental health care after emergencies. https://www.who.int/publications/i/item/9789241564571 (accessed August 31 2023)

World Health Organization (2017) mhGAP training manuals for the mhGAP intervention guide for mental, neurological and substance use disorders in non-specialized health settings, version 2.0 (for field testing) . World Health Organization. https://apps.who.int/iris/handle/10665/259161 (accessed August 31 2023).

World Health Organization (2021a) Bridging the divide: a guide to implementing the Humanitarian-Development-Peace Nexus for health. https://iris.who.int/handle/10665/351260 (accessed August 31 2023)

World Health Organization (2021b) Mental Health Atlas 2020. https://www.who.int/publications/i/item/9789240036703 (accessed 22 October 2023)

World Health Organization (2022) World mental health report. https://www.who.int/publications/i/item/9789240049338 (accessed 22 October 2023)

World Health Organization and UNICEF (2022). EQUIP - Ensuring Quality in Psychological Support. https://www.who.int/teams/mental-health-and-substance-use/treatment-care/equip-ensuring-quality-in-psychological-support (accessed 22 October 2023)

Wroe EB, McBain RK, Michaelis A, Dunbar EL, Hirschhorn LR and Cancedda C (2017) A novel scenario-based interview tool to evaluate nontechnical skills and competencies in global health delivery. Journal of Graduate Medical Education 9(4), 467–472. 10.4300/JGME-D-16-00848.1

Young T, Pakenham KI, Chapman CM and Edwards MR (2022) Predictors of mental health in aid workers: meaning, resilience, and psychological flexibility as personal resources for increased wellLbeing and reduced distress. Disasters 46(4), 974–1006. Doi: 10.1111/disa.1251

