## Supplementary material for "Identifying Core Global Mental Health Professional Competencies: A Multi-Sectoral Perspective": Multi-Sectoral Global Mental Health Employability Framework

University of Glasgow

### Multi-Sectoral Global Mental Health Employability Framework

This Multi-Sectoral Global Mental Health Employability Framework was generated from an analysis of relevant job advertisements from various job categories such as advocacy, clinical work, research, intervention development, policy, and capacity development, together with stakeholder consultations. The Framework captures a comprehensive set of in-demand skills and competencies. It has an international scope and includes a range of sectors and employers such as government agencies, NGOs, charities, higher education institutions, the private sector, and humanitarian aid organisations. The Framework is aimed at global mental health students, trainees, practitioners, trainers and educators. It is intended to aid self-reflection, career awareness, professional development and progress monitoring, and, ultimately, enhance employability in the competitive global job market.

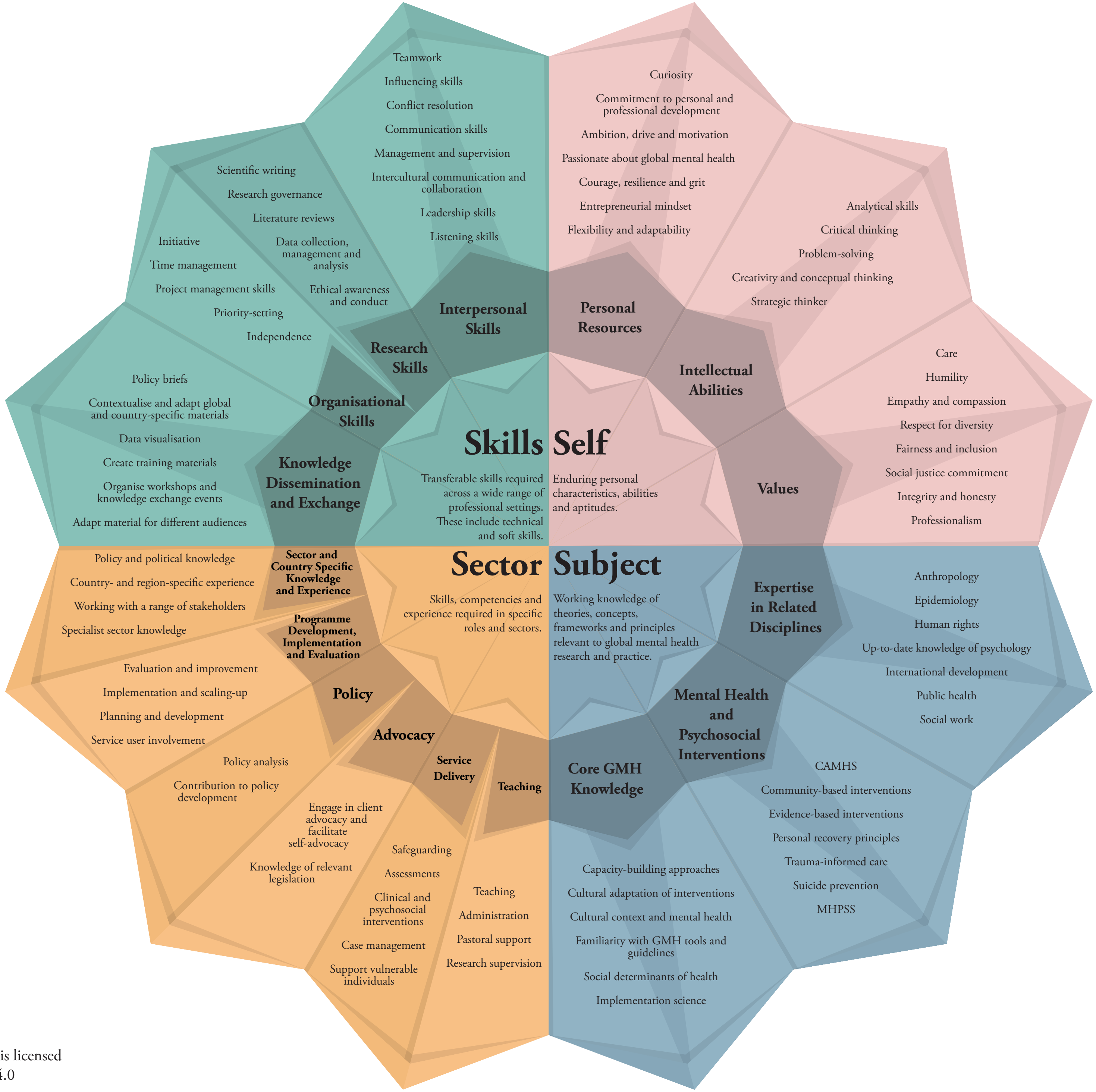


Version 1.0 of July 2023.

This work by the University of Glasgow is licensed under an Attribution-NonCommercial 4.0 International (CC BY-NC 4.0) license.
